# Computation of longitudinal phenotypes in 466 individuals with a developmental and epileptic encephalopathy enables clinical trial readiness

**DOI:** 10.1101/2023.03.02.23286645

**Authors:** Elise Brimble, Joseph Kim, Richard L. Martin, Dianalee McKnight, Alix M. B. Lacoste

**Affiliations:** Invitae, Corp, San Francisco, California, 94017, United States

## Abstract

Electronic health records (EHRs) represent a rich data source to support precision medicine, particularly in disorders with small and heterogeneous populations where longitudinal phenotypes are poorly characterized. However, the impact of EHR data is often limited by incomplete or imperfect source documentation and the inability to leverage unstructured data. Here, we address these shortcomings through a computational analysis of one of the largest cohorts of developmental and epileptic encephalopathies (DEEs), representing 466 individuals across six genetically defined conditions. The DEEs encompass debilitating pediatric-onset disorders with high unmet needs for which treatment development is ongoing. By applying a platform approach to data curation and annotation of 18 clinical data entities from comprehensive medical records, we characterize variation in longitudinal clinical journeys. Assessments of the relative enrichment of phenotypes and semantic similarity analysis highlight commonalities and differences between the six cohorts. Evaluation of medication use reflects unmet needs, particularly in the management of movement disorders. We also present a novel composite measure of seizure severity that is more robust than existing measures of seizure frequency alone. Finally, we show that the attainment of developmental outcomes, including the ability to sit independently and the ability to walk, is correlated with seizure severity scores. Overall, the combined analyses demonstrate that patient-centric real world data generation, including structuring of medical records, holds promise to improve clinical trial success in rare disorders. Applications of this approach support improved understanding of baseline disease progression, selection of relevant endpoints, and definition of inclusion and exclusion criteria.

## Introduction

The developmental and epileptic encephalopathies (DEEs) represent conditions where both epileptic activity and an underlying genetic etiology contribute to abnormal development.^1^ The promise of precision medicine for DEEs is increasing, as demonstrated through early successes in pre-clinical studies for Dravet syndrome,^2^ SCN2A encephalopathy,^3^ and Angelman syndrome,^4^ among others. With a growing population of DEEs that are candidates for targeted therapies, scalable approaches to characterization of natural history are required to define cohorts of interest, support the development of meaningful outcome measures, and optimize clinical trial design. However, this has remained challenging for the DEEs as they represent small, heterogeneous populations that are geographically distributed with variable access to clinical and research expertise. This is exacerbated in conditions where genotype-phenotype correlations have been described; ascertainment bias may contribute to underrepresentation of more moderate phenotypes, and the ability to identify phenotypic differences within a cohort can be diluted in smaller case series.^5^

The US Food and Drug Administration (FDA) defines real world data (RWD) as “data relating to patient health status and/or the delivery of health care routinely collected from a variety of sources”.^6^ Increasingly, RWD is included as part of submission packages to the FDA, particularly in those for orphan products given its ability to address limitations in traditional study design.^7, 8^ Between 2019 through the first half of 2021, 85% of submissions to the FDA included RWD.^7^ With high healthcare utilization established for individuals living with a rare diagnosis,^9^ electronic health records (EHRs) represent a comprehensive and longitudinal RWD source that captures both the onset of symptoms and their evolution. However, the value of this data source is dependent on the ability to address and accommodate for key limitations, including fragmentation of healthcare; variability in documentation practices; and ability to leverage unstructured text, which represents >80% of EHR content.^10^

More recently, the application of ontologies has emerged as a viable strategy to harmonize disparate data sources where inconsistent terminology and specificity is used to describe clinical findings. Ontologies represent structured, hierarchical frameworks that allow for characterization of data, such as phenotypes, through computational methods.^11^ Through the use of ontologies, narrative data sources can be structured by applying a controlled vocabulary with established relationships between increasingly specific concepts. This approach has been successfully applied to characterize the epilepsy phenotype of children cared for at a large pediatric epilepsy center^12, 13^ as well as in genetic DEEs,^14–17^ to establish genotype-phenotype correlations,^15, 17^ and to expedite genetic diagnosis.^18^ The use of longitudinal EHR data combined with mapping to standard ontologies provides unprecedented breadth and depth of medical history data for representative samples of rare populations. However, this approach can be further strengthened by expanding data capture beyond individual health systems and through use of a common framework for data generation and evaluation.

In this study, we describe the use of a novel patient-centric RWD platform to establish robust characterization of six DEEs, representing 466 individuals with causative variants in: *FOXG1* (n=90; MIM: 613454), *KCNQ2* (n=33; MIM: 613720), *SCN2A* (n=49; MIM: 613721), *SCN8A* (n=67; MIM: 614558), *STXBP1* (n=85; MIM: 612164), and *SYNGAP1* (n=142; MIM: 612621). In aggregate, we reviewed the equivalent of >3,000 patient years of data through evaluation of 97,256 annotated clinical concepts. We employed computational analyses of phenotypes, medications and physical exam findings mapped to an ontology to illustrate respective patient journeys, as well as intra- and inter-cohort similarity. We also present a novel composite measure of seizure severity to characterize the epilepsy phenotype longitudinally and correlate with developmental outcomes.

## Subjects and Methods

### Study Participants

The study population was drawn from individuals enrolled in Ciitizen®, a wholly owned subsidiary of Invitae Corporation. Although the platform is made available to any individual with a rare neurodevelopmental disorder, the majority of enrollment is achieved through engagement and collaboration with patient advocacy groups. Genetic diagnoses were verified through collection and review of clinical genetic test reports. All variants were reviewed and classified as pathogenic, likely pathogenic, or a variant of uncertain significance (VUS) in accordance with guidance outlined by the American College of Medical Genetics and Genomics; individuals with likely benign or benign variants were excluded from analyses.^19^ Individuals with VUS were included if at least two of the following criteria were met: (1) the variant was *de novo* with assumed parentage, (2) the ordering physician considered the VUS to be diagnostic and managed the participant accordingly, and/or (3) internal review demonstrated a consistent phenotype for the associated DEE. Individuals were eligible to participate if they had a qualifying variant in any of the six genes of interest: *FOXG1, KCNQ2, SCN2A, SCN8A, STXBP1* or *SYNGAP1*. Individuals with a second genetic diagnosis associated with an overlapping clinical phenotype were excluded. Similarly, individuals with copy number variants encompassing more than one of the six genes above were excluded.

Caregivers and/or legal guardians of study participants provided broad consent to share de-identified data for research. The generation and subsequent analysis of participant data received determinations of exemption through a central IRB.

### Generation of RWD through the Ciitizen Platform

Ciitizen is a patient-centric RWD platform that transforms medical records into structured datasets that describe phenotypes and medical interventions at the request of patient participants.^20^ Ciitizen has developed an end-to-end operation that spans medical record collection through to data delivery. Medical records were collected from institutions within the United States where participants received care using the right of access granted by the Health Insurance Portability and Accountability Act (HIPAA). Following record retrieval, each participant was reviewed for completeness by trained clinicians; only those individuals with sufficient and specific documentation surpassing defined minimum thresholds were included. Example completeness criteria include collection of the participant’s genetic test report, as well as at least annual documentation by relevant physicians and clinicians.

Ciitizen has developed tooling to scale data harmonization and organization from unstructured medical record documents, including physician notes, diagnostic imaging reports, and genetic test reports. In brief, each document was systematically evaluated for relevant data variables, including clinical phenotypes and diagnoses, medication use, therapeutic procedures, hospitalizations, attainment of developmental milestones, and seizure severity. In aggregate, scope of data capture included 18 unique clinical data entities, encompassing 48 variable fields (**Table S1**). To correct for variation in clinical documentation, extracted data were mapped to an internationally recognized terminology;^21^ diagnoses, observations, and procedures were mapped to SNOMED CT (US Edition, version 2022_03_01),^22^ whereas medications were mapped to RxNorm (version 20AA_220307F).^23^ Throughout the text, annotated concepts are presented as “clinical concept, SNOMED CT code” to provide documentation of both the assigned name and static code. In annotation, the most specific concepts were used to ensure fidelity of data capture. Resulting data underwent de-duplication through a process of aggregation, where like data variables are condensed into a representative “entity” that spans a single date or date range. The resulting concepts can be compared within and across cohorts, longitudinally. Extracted data undergo review by a team of advanced practice providers with relevant training and clinical experience who confirm accurate annotation and perform source document verification. The resulting data elements were stored securely in a HIPAA-compliant, controlled access, indexed database.

### Enrichment Analysis and Terminology Propagation

To support clinical data analysis across individuals, we leveraged the hierarchical structure of SNOMED CT to propagate annotated concepts to more general “ancestral” concepts. To reflect specificity of cohort data, the highest-level ancestral concepts used throughout propagation were limited to those that were recorded for at least one participant in the sample dataset. For example, the clinical concept of “absence seizure, 79631006” would be mapped up to an ancestral concept of “seizure, 91175000” by way of the increasingly non-specific concepts “generalized onset seizure, 246545002” and “epileptic seizure, 313307000”. However, propagation would not proceed to “finding of brain, 299718000” as this ancestral concept was not documented for any patient in the sample dataset. As such, we enabled a degree of concept normalization to facilitate comparisons amongst individuals, while avoiding the use of excessively nonspecific clinical concepts.

To establish “enrichment” of clinical diagnoses and observations, including physical exam findings, we compared frequency of a given annotated or ancestral concept in one cohort to its frequency in the remaining cohorts. Concepts were considered to be enriched if their frequency in one cohort was ≥2.5-fold than in the remaining data set. Only concepts present in at least 10 individuals in the broader cohort were included in this analysis.

### Intra- and Inter-cohort Phenotypic Similarity

To evaluate phenotypic similarity between DEE cohorts, we leveraged the ontology of SNOMED CT to calculate information content (IC) for the most informative common ancestor (mica) of two given SNOMED CT codes. This approach has been described previously,^24, 25^ and employed in other DEE cohorts using ontological annotation.^14–17^ We computed the IC for each concept, defined as -log_2_(*f*), where *f* is the frequency of the term in the sampled population. Therefore, greater IC values correspond to increasing specificity. For example, general concepts common to most participants, such as “developmental disorder, 5294002” (IC = 0.00) and “seizure, 91175000” (IC = 0.20) have the lowest computed IC. In contrast, clinical concepts associated with increased specificity, like “right hemiplegia, 278284007” (IC = 6.14) and “vocal tic disorder, 230336005” (IC = 6.14) have the highest IC. With propagation of individual concepts, as described above, the most informative common ancestor represents a shared ancestor between two concepts with the highest IC score (sim_mica). To represent a similarity score for a cohort of individuals, we calculated mean sim_mica scores for 2000 randomly sampled participant pairs.

### Evaluation of Epilepsy Severity using a Novel Composite Score

To quantify relative burden of epilepsy, we developed a composite seizure severity score that encompasses four domains: (i) seizure frequency, (ii) use of concomitant anti-seizure medications (ASMs), (iii) frequency of prolonged seizures (>5 minutes), and (iv) frequency of hospitalizations for prolonged seizures or increased seizure frequency. In doing so, we sought to better reflect disease burden beyond seizure frequency alone. For each domain, individuals were scored on a 6-point scale, with the exception of frequency of prolonged seizures which was scored on a 3-point scale (**Table S2**). To illustrate the progression of seizure burden across cohorts, each domain was evaluated in yearly intervals, with the maximum score recorded in that year contributing to the representative composite score. Analyses were restricted to ages 0 through 16 given reduced sample sizes at older ages. Higher composite seizure severity scores are associated with greater seizure burden.

Performance of the composite score was compared to a previously reported scale used in the evaluation of relative seizure burden in a pediatric cohort as documented in medical records.^26^ To evaluate sensitivity and robustness of the composite score to data omission, we measured the percent change in score as a result of random deletion of 10% or 20% of the source data.

### Statistical Analyses

Computations were performed using Graphpad Prism 9 and Python framework with scipy, statsmodel, and seaborn packages for statistical analysis and production of visualizations. When plotting central tendency measures such as average seizure severity scores over time, 95% confidence intervals are presented. For mean comparisons, nonparametric statistical tests were performed at significance level of 0.05 with Bonferroni correction for multiple comparisons. An Ordinary Least Square (OLS) regression model was fit when investigating the relationship between seizure severity scores and the time for patients to be able to perform development milestone tasks.

## Results

### Computational analysis of structured medical record data can generate detailed patient journeys

Data from 466 individuals with a DEE enrolled in the Ciitizen platform were analyzed, representing participants with causative genetic variants in one of six genes: *FOXG1, KCNQ2, SCN2A, SCN8A, STXBP1,* and *SYNGAP1* (**Table 1, Figure S1**).

**Table 1.** Demographic information for six DEE cohorts

| Cohort | n | male (%) | female (%) | Mean age<br>± SD | # unique concepts |  |  |
| --- | --- | --- | --- | --- | --- | --- | --- |
|  |  |  |  |  | diagnosis | exam findings | seizure type |
| *FOXG1* | 90 | 57 (63.3) | 33 (36.7) | 7.5 ± 5.4 | 438 | 216 | 30 |
| *KCNQ2* | 33 | 17 (51.5) | 16 (48.5) | 7.4 ± 5.7 | 381 | 190 | 28 |
| *SCN2A* | 49 | 25 (51.0) | 24 (49.0) | 7.2 ± 4.9 | 381 | 189 | 31 |
| *SCN8A* | 67 | 33 (49.3) | 34 (50.7) | 8.4 ± 5.6 | 465 | 217 | 34 |
| *STXBP1* | 85 | 41 (48.2) | 44 (51.8) | 8.3 ± 6.7 | 380 | 201 | 27 |
| *SYNGAP1* | 142 | 72 (50.7) | 70 (49.3) | 9.1 ± 7.8 | 449 | 175 | 25 |
| **All** | **466** | **245 (52.6)** | **221 (47.4)** | **8.2 ± 6.5** | **798** | **364** | **45** |

Demographic variables are presented for all participants, including cohort size, sex assigned at birth, and age at analysis. In addition, the total number of unique annotated clinical concepts are shown for diagnoses, physical exam findings, and seizure types.

There were no significant differences in age or biological sex between cohorts. The average age of participants at the time of analysis was 8.2 years (range 0.7 - 65.9), and average age at diagnosis was 3.0 years (range 0.04 - 12.7). Age at diagnosis was significantly lower for individuals with a variant in *KCNQ2* compared to all cohorts except *SCN2A*; conversely, age at diagnosis was significantly higher in individuals with a variant in *SYNGAP1* compared to all cohorts except *SCN8A* (**Figure S2A**). The correlation between age at diagnosis and age at seizure onset ranged from no correlation (R^2^ = 0.0000, *KCNQ2*) to a moderate correlation (R^2^ = 0.4142, *SCN2A*) (**Figure S2B**). High comprehensiveness of medical record collection was demonstrated through the number of unique institutions (mean 5.84 per participant, range 1-30), the volume of medical records (mean 1211.00 pages per participant, range 33-16,972) and representative years of patient data (mean 7.77 years per participant, range 1-48) (**Table S3**). The number of years of data represented suggests that the resulting dataset extends to the time of symptom onset. In aggregate, 3,424 patient years of data were reviewed. The average duration from first enrolled participant through to data generation for all participants in a cohort was 1.39 years ± 0.43 SD across the individual cohorts described here.

Clinical phenotypes described in participant medical records were annotated with 798 unique and 97,256 total SNOMED CT concepts to support harmonization across disparate data sources. To account for variability in specificity of documentation, assigned concepts were also propagated to the most general ancestral concept used at least once in the DEE cohort (see **Methods**). We evaluated relative frequency of ancestral concepts present in at least half of one or more cohorts (**Table 2**). The most commonly documented ancestral concepts across all cohorts were: “developmental disorder, 5294002”; “seizure, 91175000”; “altered bowel function, 88111009”; “hypotonia, 398152000”; and “gastroesophageal reflux disease, 235595009”. Even amongst these “common” phenotypes, we observed emerging cohort-specific patterns. For example, the ancestral concepts “movement disorder, 60342002”, “communication disorder, 278919001”, and “gastroesophageal reflux disease, 235595009” were documented at significantly lower frequencies in individuals with *SYNGAP1* variants compared to the remaining cohorts; similarly, “dependency on feeding tube, 12991000224107” was observed at significantly lower frequencies in those with *STXBP1* or *SYNGAP1* variants. We also observed that “seizure free, 370994008” was documented at significantly higher frequencies in individuals with a *KCNQ2*-related disorder, reflecting established disease trajectory.^27^

**Table 2.** Relative frequency of common ancestral phenotypes

| phenotype | Code | *FOXP1* | *KCNQ2* | *SCN2A* | *SCN8A* | *STXBP1* | *SYNGAP1* |
| --- | --- | --- | --- | --- | --- | --- | --- |
| developmental disorder | 5294002 | 0.989* | 0.970 | 1.000 | 1.000 | 1.000 | 1.000 |
| microcephaly | 1148757008 | 0.878* | 0.091 | 0.122 | 0.179 | 0.047 | 0.049 |
| altered bowel function | 88111009 | 0.833* | 0.727 | 0.735 | 0.731 | 0.518 | 0.599 |
| seizure | 91175000 | 0.744 | 1.000* | 0.755 | 0.940* | 0.741 | 0.831 |
| hypotonia | 398152000 | 0.733 | 0.545 | 0.755 | 0.642 | 0.800* | 0.514 |
| movement disorder | 60342002 | 0.722 | 0.576 | 0.490 | 0.552 | 0.471 | 0.225* |
| strabismus | 22066006 | 0.722* | 0.545* | 0.306 | 0.254 | 0.153 | 0.211 |
| gastroesophageal reflux disease | 235595009 | 0.678 | 0.576 | 0.673 | 0.552 | 0.400 | 0.232* |
| dysphagia | 40739000 | 0.656* | 0.485 | 0.388 | 0.433* | 0.200 | 0.261 |
| feeding problem | 78164000 | 0.633* | 0.606 | 0.551 | 0.448 | 0.306 | 0.408 |
| visual disturbance | 63102001 | 0.600* | 0.394 | 0.347 | 0.284 | 0.176 | 0.113* |
| sialorrhea | 53827007 | 0.500 | 0.545 | 0.408 | 0.403 | 0.247 | 0.317 |
| dependency on feeding tube | 12991000224107 | 0.489 | 0.636 | 0.408 | 0.433 | 0.106* | 0.056* |
| increased muscle tone | 56731001 | 0.478 | 0.545 | 0.286 | 0.299 | 0.271 | 0.127* |
| communication disorder | 278919001 | 0.167 | 0.152 | 0.327 | 0.224 | 0.318 | 0.585* |
| seizure free | 370994008 | 0.144 | 0.606* | 0.020 | 0.075 | 0.094 | 0.120 |
| tremor | 26079004 | 0.056 | 0.121 | 0.122 | 0.343* | 0.565* | 0.134 |

Relative frequency of ancestral clinical concepts present in $\geq 50\%$ of one of the six DEE cohorts are shown. Values with asterisks represent those that are significantly different from one or more cohorts.

The Ciitizen platform enables collection of longitudinal data sourced from medical records, allowing for detailed mapping of symptom onset and evolution with disease progression. We visualized age at onset for the common ancestral clinical concepts highlighted above, as well as age of genetic diagnosis (**Figure 1**). In ordering ancestral concepts by median age of onset, cohort-specific sequencing of phenotypes becomes apparent. For cohorts *KCNQ2, SCN2A, SCN8A,* and *STXBP1*, the presenting ancestral clinical concept for most participants was “seizure, 91175000”, with onset in early infancy. Conversely, median age at seizure onset for individuals with variants in *SYNGAP1* was in early childhood (median = 2.71 years). “Feeding problem, 78164000” and “gastroesophageal reflux disease, 235595009” were other common presenting diagnoses, with “dependency on feeding tube, 12991000224107’’ documented early for *KCNQ2*. In aggregate, by mapping the patient journey in this manner, we effectively illustrate high disease burden in early childhood for the six DEEs evaluated.

**Figure 1.**
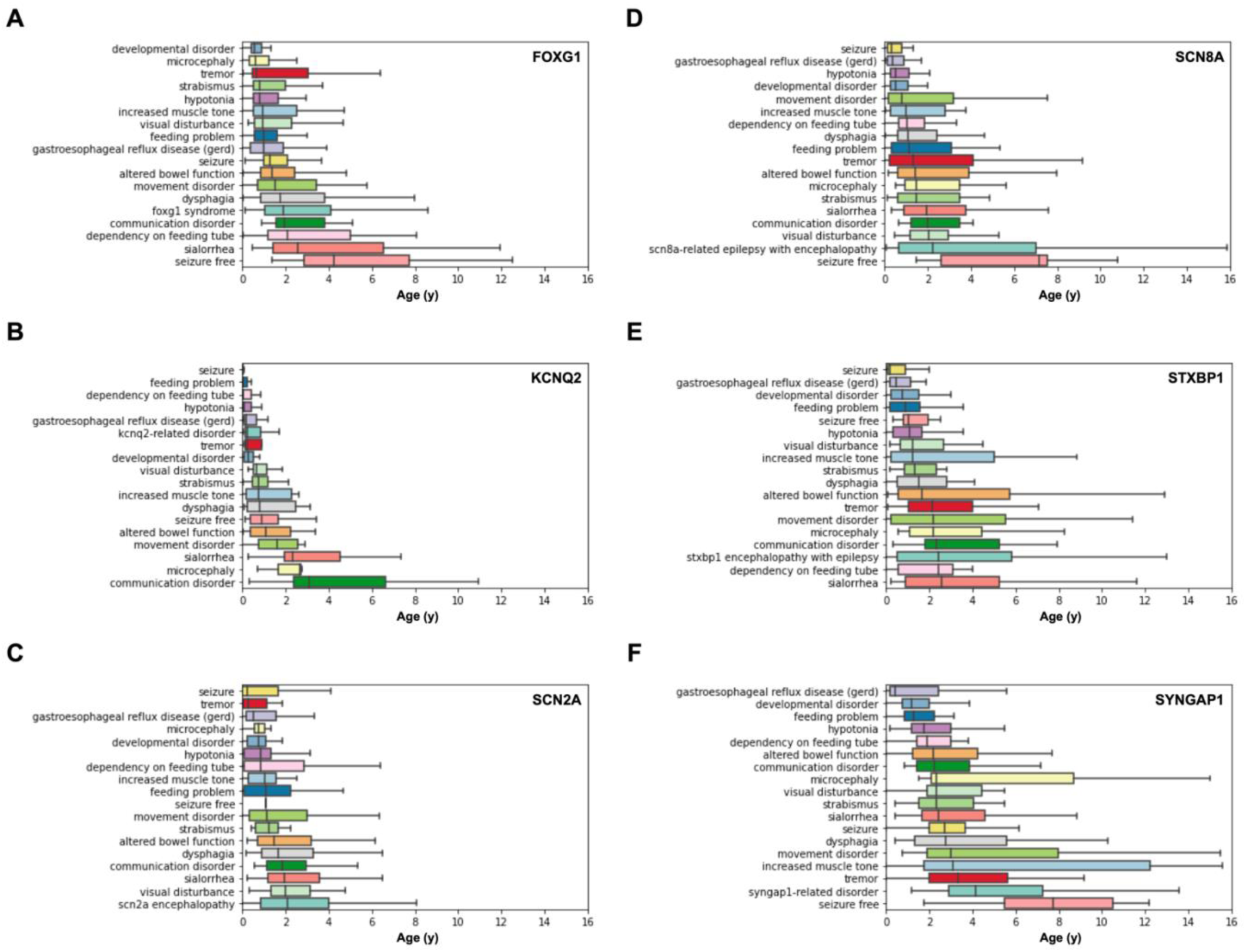
Age of onset of common ancestral clinical concepts across the six DEE cohorts. To illustrate respective patient journeys, age of onset for phenotypes present in ≥50% of one of the cohorts are shown for *FOXG1* **(A)**, *KCNQ2* **(B)**, *SCN2A* **(C)**, *SCN8A* **(D)**, *STXBP1* **(E)**, and *SYNGAP1* **(F)**. Individual concepts have been propagated to the least specific ancestral concept present in the cohort. Boxes indicate the interquartile range (IQR), including the median line, by which phenotypes are sorted. Whiskers represent remaining data points up to a maximum length of 1.5x IQR; outliers beyond this point are not presented.

### Relative enrichment of clinical phenotypes, physical exam findings, and seizure types reliably discriminates between DEE cohorts

With a growing number of precision therapies available for genetic epilepsies, early diagnosis is critical in maximizing treatment success.^28^ We identified clinical phenotypes enriched at least 2.5-fold in each DEE cohort to support differentiation in the first year of life (**Table 3**). To leverage the specificity afforded by annotation, only non-propagated SNOMED CT concepts were analyzed. As illustrated in **Figure 1**, presence of “epilepsy, 84757009” and/or “neonatal seizures, 87476004” was enriched in three cohorts in infancy: *KCNQ2*, *SCN8A*, and *STXBP1*. In addition, presence of “infantile spasms, 723437000” was enriched in individuals with variants in *KCNQ2*. The *FOXG1* cohort was differentiated at this early time point by enrichment of “microcephaly, 1148757008”, “strabismus, 22066006”, “cortical visual impairment, 413924001”, and “chronic constipation, 236069009”.

**Table 3.** X-fold enrichment of clinical phenotypes in the first year of life

| diagnosis | Code | *FOXP1* | *KCNQ2* | *SCN2A* | *SCN8A* | *STXBP1* | *SYNGAP1* |
| --- | --- | --- | --- | --- | --- | --- | --- |
| microcephaly | 1148757008 | 23.396 |  |  |  |  |  |
| epilepsy | 84757009 |  | 2.952 |  | 2.799 |  |  |
| feeding difficulty | 299698007 |  | 2.708 |  |  |  |  |
| dependency on feeding tube | 12991000224107 |  | 6.029 |  |  |  |  |
| esotropia | 16596007 | 5.670 |  |  |  |  |  |
| failure to thrive | 54840006 | 3.064 |  |  |  |  |  |
| cortical visual impairment | 413924001 | 4.642 |  |  |  |  |  |
| chronic constipation | 236069009 | 2.639 |  |  |  |  |  |
| strabismus | 22066006 | 6.963 |  |  |  |  |  |
| appendicular hypertonia | 13291000224105 |  | 5.047 |  |  |  |  |
| decreased muscle tone | 398151007 |  | 4.977 |  | 2.552 |  |  |
| dependence on supplemental oxygen | 931000119107 |  | 6.298 |  |  |  |  |
| infantile spasms | 723437000 |  | 2.678 |  |  |  |  |
| apnea | 1023001 |  |  |  | 6.451 |  |  |
| tremor | 26079004 |  |  |  |  | 2.637 |  |
| neonatal seizures | 87476004 |  |  |  |  | 5.603 |  |

Clinical concepts shown here represent those enriched $\geq 2.5$ -fold in at least one of the six DEE cohorts, during the first year of life.

Both *SCN8A* and *KCNQ2* were associated with respiratory challenges, with “apnea, 1023001” and “dependence on supplemental oxygen, 931000119107” enriched, respectively. Although similar trends persisted throughout the first six years of life, further differentiation was noted to reflect onset or physician recognition of symptoms in childhood (**Table S4**). Notably, emergence of the previously described hyperkinetic-dyskinetic movement disorder associated with FOXG1 syndrome^29–31^ was demonstrated by enrichment of concepts “dystonia, 15802004”, “chorea, 271700006”, and “choreoathetosis, 43105007”. Characteristic features of SYNGAP1-related disorder are also enriched for during this time, including concepts related to neuropsychological features (“aggressive behavior, 61372001”; “autism suspected, 401204006”; and “sensory integration disorder, 425988004’’) as well as “generalized epilepsy, 19598007”. These clinical features may also represent viable endpoints for use in clinical trials.

We performed a similar assessment of physical exam findings within the first six years of life (**Table 4**). The number of documented neurology exams ranged from an average of 4.5 (*SYNGAP1*) to 15.0 (*SCN8A*) per participant, representing a total of 3,458 reviewed exams. Individuals with variants in *FOXG1* had the greatest number of enriched exam findings, and those findings were consistent with observed clinical phenotypes of strabismus, a hyperkinetic movement disorder, and limited functional hand use. Further, enrichment for the combination of “truncal hypotonia, 13421000224108” and “appendicular hypertonia, 13291000224105” was observed; this pattern emerged in the first year of life and persisted through the first six years. Physical exam findings associated with impaired coordination were notably enriched for in those with *STXBP1* variants, including “dysmetria, 32566006”, “titubation, 14981000224100”, “ataxia, 20262006”, “tremor, 26079004”, and “intention tremor, 30721006”. Concepts describing gait abnormalities were enriched for in the *SYNGAP1* cohort, including “abnormal gait, 22325002” and “unsteady gait, 22631008”.

**Table 4.**
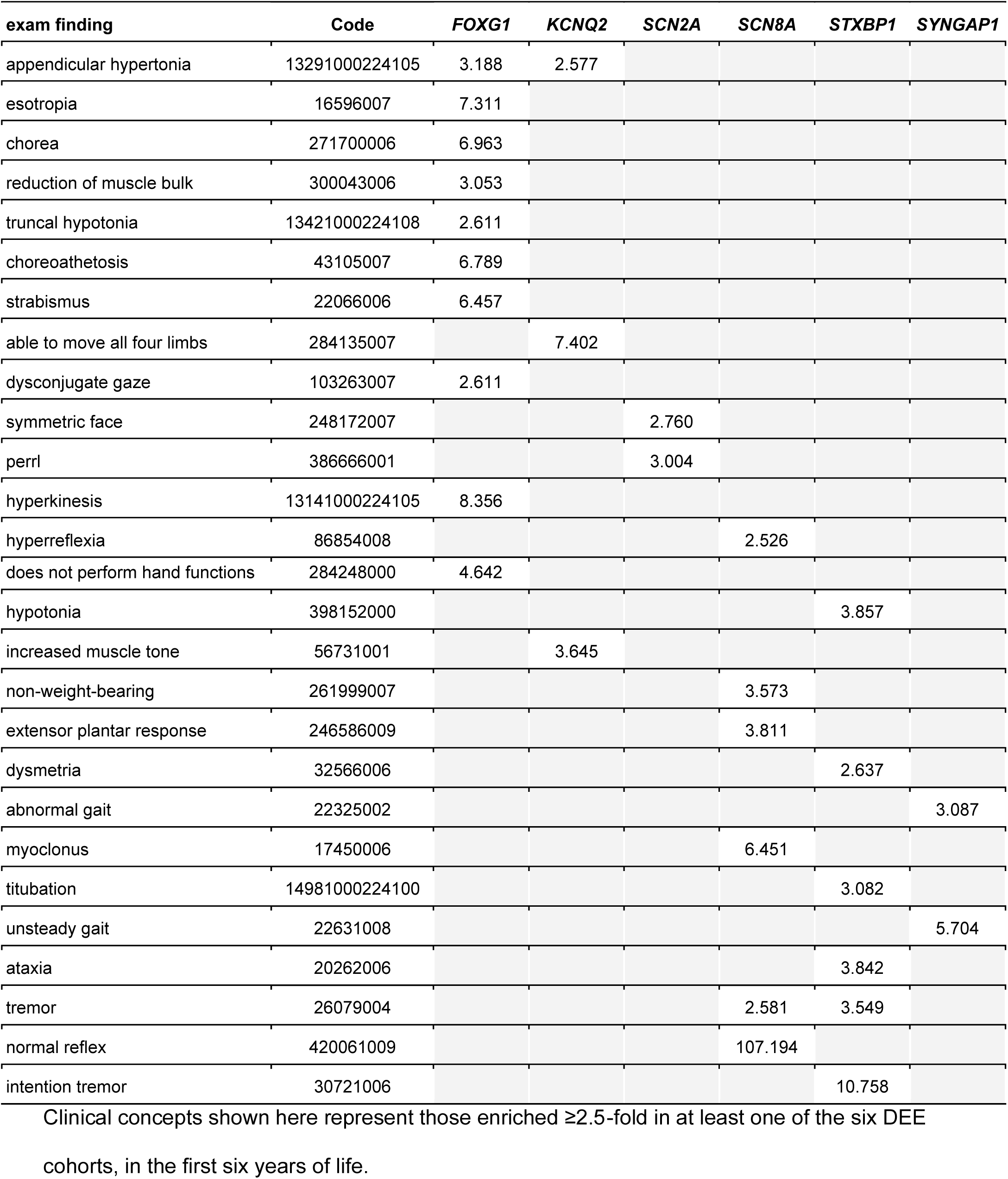
X-fold enrichment of physical exam finding concepts in the first six years of life

### Annotated medication use reflects onset and evolution of clinical phenotypes

To complement and validate clinical phenotype mapping, we reviewed annotated medication use and the indication for use within the six cohorts. Individual medications were mapped to one of 11 indications, including behavioral differences, gastrointestinal problems, movement disorder, neuropsychological diagnoses, seizures, and sleep (**Figure 2**). As would be expected, we observed expansion in the proportion of medications used for a given indication with expected age of onset for the target symptom. For example, in the *SYNGAP1* cohort, the relative proportion of medications used for a neuropsychological indication or seizures increases outside of infancy. Conversely, use of medications for seizure management represents a significant relative proportion of medications documented early in life for *KCNQ2* and *SCN8A*. The observed patterns in medication use are consistent with the emergence of their associated symptoms as documented by clinical phenotype annotation. With age, the indication for medication use remains relatively stable.

**Figure 2.**
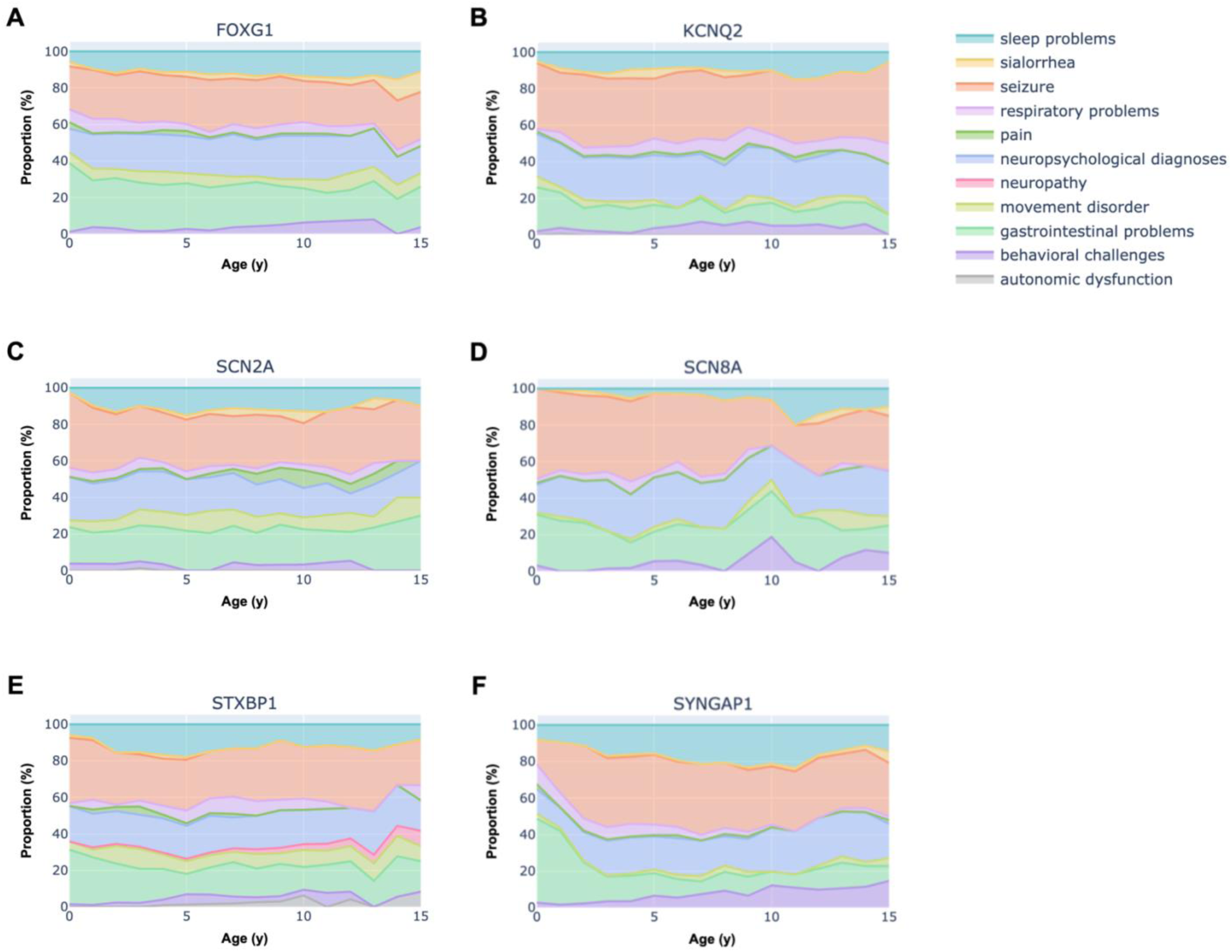
Relative frequency of indications for medicinal intervention. Relative proportion of medication use for common indications is shown for *FOXG1* **(A)***, KCNQ2* **(B)***, SCN2A* **(C)***, SCN8A* **(D),** *STXBP1* **(E)***, SYNGAP1* **(F)** across ages 0-15 years. Each individual medication was mapped to one of 11 indications. The percentage denoted on the y-axis reflects the proportion of medications grouped by indication, for each year of age assessed. To determine whether specific seizure types were more common in any of the six DEE cohorts, we performed enrichment analysis for annotated seizures in the first six years of life. We observed enrichment of “prolonged seizure (>5 minutes), 13961000224109” and “status epilepticus, 230456007” in the *SCN8A* cohort. For individuals with a *SYNGAP1* variant, concepts representing generalized seizures were enriched for, including: “absence seizure, 79631006”, “atypical absence seizure, 23374007”, and “atonic seizure, 42365007”. No specific seizure types were enriched for in the other DEE cohorts, reflecting both overlap and variability in the associated epilepsy phenotypes.

We observed that clinical phenotypes and physical exam findings describing features of a movement disorder were enriched for in participants with variants in *FOXG1* and *STXBP1*. However, the documentation of medications for this indication is discordant, demonstrating relatively limited medical management. As a prominent phenotype that impacts function, we highlight treatment of movement disorders as an unmet need in these two populations.

### Greatest intra- and inter-cohort phenotypic similarity observed for *FOXG1* and *KCNQ2*

Semantic similarity algorithms can be used to measure the degree of similarity between two individuals or cohorts of interest. To characterize phenotypic heterogeneity, we first sought to understand how similar individuals with the same genetic diagnoses (i.e., intracohort) were to each other at <1 year (**Figure 3A**), <3 years (**Figure 3B**), and <6 years (**Figure 3C**). The greatest intracohort similarity (high sim_mica scores) was observed in individuals with a variant in *FOXG1*, emerging at <3 years and persisting. This suggests that early in life, FOXG1 syndrome is associated with a recognizable phenotypic gestalt that distinguishes it from the other DEEs studied here. In contrast, more limited phenotypic similarity was observed in the *STXBP1* and *SYNGAP1* cohorts at all three timepoints. In fact, the majority of individuals with *SYNGAP1* variants show a similarity score that is close to zero when compared to others in that cohort at <1 year. This is consistent with the high variability in onset of phenotypes shown in **Figures 1E** and **1F**. Over time, individuals within the same genetic cohort appear more similar to each other, reflecting accumulation of shared phenotypes with age (**Figure 3D**).

**Figure 3.**
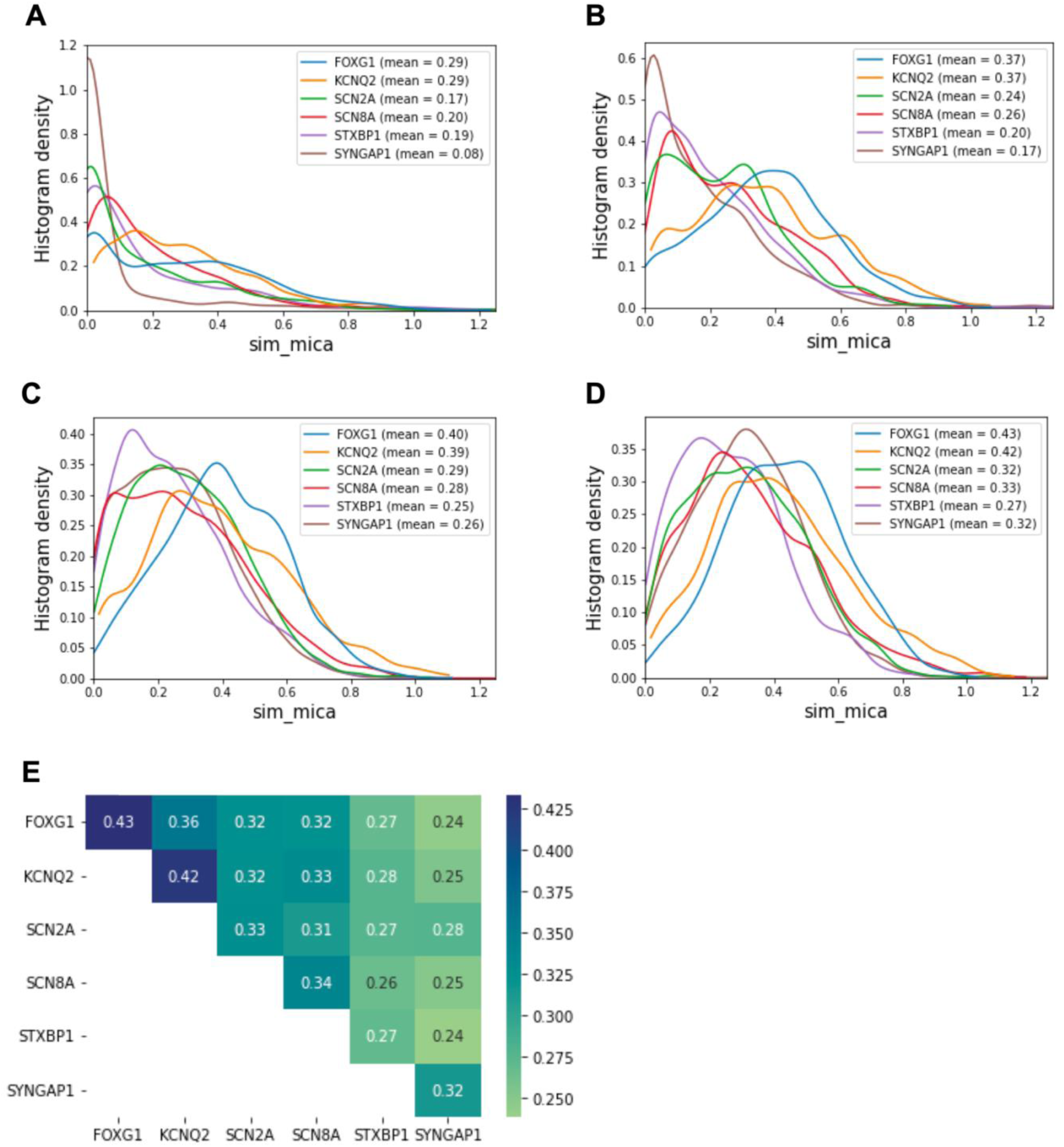
Evaluation of intra- and inter-cohort phenotypic similarity. To quantify phenotypic similarity amongst individuals within DEE cohorts, we employed a semantic similarity algorithm and showed the distribution of similarity scores at three timepoints: <1 year **(A)**, <3 years **(B)**, and <6 years **(C)**. Average similarity score by year of age is shown in **(D)**. Higher scores indicate greater intracohort similarity. To evaluate intercohort phenotypic similarity, the same semantic similarity algorithm was performed with permutations of paired patients from distinct cohorts **(E)**. Greater sim_max scores are associated with increased similarity.

We applied the same approach to determine similarity between DEE cohorts (i.e., intercohort). Greatest phenotypic similarity was observed between the *FOXG1* and *KCNQ2* cohorts (**Figure 3E**). Conversely, the lowest phenotypic similarity between cohorts was observed for *STXBP1* and *SYNGAP1*. Both cohorts also demonstrated more limited phenotypic similarity to the remaining four cohorts, which was most notable for *SYNGAP1*. Of the other five DEE cohorts, the greatest phenotypic similarity for the *SYNGAP1* cohort was observed with *SCN2A*. This may reflect the relative enrichment in both cohorts for concepts related to neuropsychological diagnoses, including suspicion for autism spectrum disorder and sensory integration disorder.

### A novel composite metric differentiates longitudinal epilepsy severity

Reduction in seizure frequency is a common primary endpoint for interventional trials in genetic epilepsies. However, evolution of the epilepsy phenotype has not been well characterized for many DEEs. This poses challenges for optimizing clinical trial design, particularly in establishing inclusion and exclusion criteria. Further, restricting an endpoint to seizure frequency may minimize the negative impact of additional variables, including adverse effects associated with anti-seizure medications (ASMs) and hospitalizations. To address these limitations, we developed a composite measure of epilepsy burden to characterize the epilepsy phenotype in FOXG1-, KCNQ2-, SCN2A-, SCN8A-, STXBP1-, and SYNGAP1-related disorders. In addition to seizure frequency, which is often considered in isolation, the composite measure also takes into account the number of concurrent ASMs used, incidence of prolonged seizures, and the number of hospital admissions for poor seizure control (**Table S2**). Compared to evaluation of seizure frequency only using a score developed by Fitzgerald *et al.*,^26^ we observed that this composite measure allowed for broader distribution of scores between cohorts (**Figures S3A, S3B**), as well as improved robustness to change in scores over time (**Figures S3C, S3D**). Expanding the range and contributors to the composite seizure severity score allowed for increased sensitivity to data omissions (**Table S5**), with relatively smaller magnitudes of change in scores (**Tables S5**).

Across the six cohorts, average seizure severity scores ranged from 5.7 ± 3.7 SD in *SYNGAP1* to 11.8 ± 4.7 SD in *SCN8A* (**Figure S3A**). The distribution of seizure severity scores by year of age is visualized as a heatmap to characterize evolution of the epilepsy phenotype over time (**Figure 4**). Consistent with findings from the enrichment analysis of clinical diagnoses, high seizure severity scores are observed in the first year of life for *KCNQ2*, *SCN8A*, and *STXBP1*. Notably, no individuals with a *KCNQ2* variant were assigned a score of 0 in the first year of life. Rapid improvement in seizure severity scores for this cohort are observed after the first year of life, again consistent with the epilepsy phenotype described for *KCNQ2*^27^ and findings from our enrichment analysis. Similarly, most individuals with a *SYNGAP1* variant for whom a score can be determined have low seizure severity scores in the first two years of life, consistent with the median age of seizure onset at 2.7 years. Interestingly, a bimodal distribution of seizure severity is observed for a subset of cohorts in the first year of life, most prominent in the *SCN2A* and *STXBP1* cohorts. This finding may reflect established genotype-phenotype correlations,^32^ or highlights opportunities to identify novel contributors to variability in the epilepsy phenotype.

**Figure 4.**
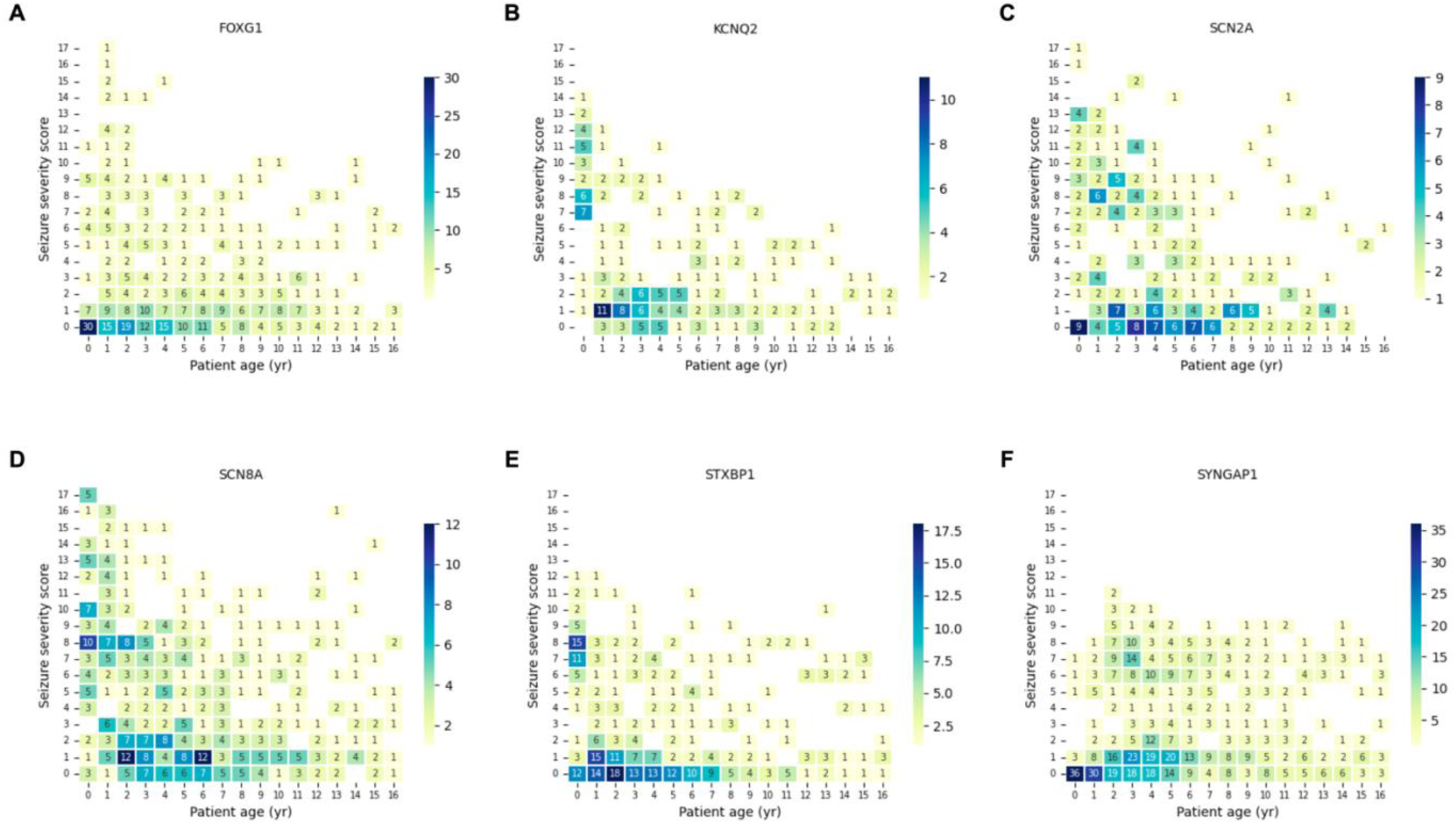
Longitudinal characterization of epilepsy severity using a composite measure. The distribution of seizure severity scores is shown by year of age for *FOXG1* **(A)**, *KCNQ2* **(B)**, *SCN2A* **(C)**, *SCN8A* **(D)**, *STXBP1* **(E)**, and *SYNGAP1* **(F)**. The color scale represents the number of individuals documented with a given seizure severity score at the noted age. Higher seizure severity scores correspond to greater burden of disease, as measured by seizure frequency, concomitant use of ASMs, hospitalizations for epilepsy, and occurrence of prolonged seizures.

### Attainment of developmental milestones is negatively correlated with seizure severity

The DEEs are in part defined by the contribution of epileptic activity to atypical development, beyond what would be expected of the genetic etiology alone.^1^ For all cohorts, we documented whether a participant was described as having achieved a developmental milestone, as well as the age of attainment, where relevant. We limited analyses to developmental milestones commonly referenced in medical records as part of the developmental history, including ability to sit independently, walk independently, and use at least one word. We grouped individual composite seizure scores into one of three severities: (i) low (score = 0-5), (ii) medium (score = 6-11), and (iii) high (score = 12-17) (**Figure 5**). The score used to assign a grouping represents the maximum score for the year the specified milestone was achieved. Greatest attainment of developmental milestones was observed in those categorized with low epilepsy severity. With increasing epilepsy burden, we observed corresponding reduction in the frequency of milestone attainment as well as increased time to milestone attainment. A consistent trend was observed when individual gene cohorts were stratified as well. The gross motor milestones, “ability to walk” and “ability to sit unsupported” were most prominently affected.

**Figure 5.**
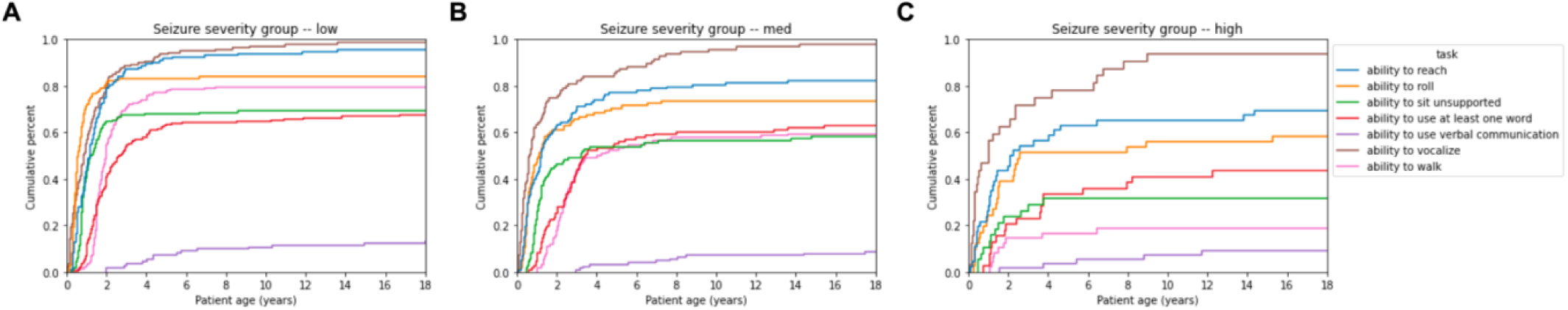
Epilepsy severity is associated with developmental outcomes. The proportion and rate of attainment for commonly reported developmental milestones is compared in individuals with low (**A,** scores 0-5), medium (**B**, scores 6-11), or high (**C**, scores 12-17) seizure severity scores.

To evaluate the contribution of seizure severity score to the predicted age of milestone attainment, we applied an OLS regression model that included the gene cohort and the specific milestone as covariates. The resulting correlation coefficient was indicative of a weak fit (R^2^ = 0.396). Therefore, seizure severity score explains some, but not all, of the variability observed in milestone attainment.

## Discussion

In this study, we demonstrated the utility of RWD in constructing robust phenotypic histories that describe disease course in detail. As access to genetic testing grows, novel solutions are required to ensure clinical characterization keeps pace. This is particularly true for early-onset epilepsies where high diagnostic yields for clinical genetic testing are well-described.^33, 34^ For example, despite its discovery as a disease-associated gene almost 20 years ago,^35^ literature describing the phenotype associated with *FOXG1* is largely limited to a handful of cross-sectional case series.^29, 31, 36, 37^ In using a novel platform for rare disorders, we describe and compare the natural history for individuals with variants in *FOXG1*, *KCNQ2*, *SCN2A*, *SCN8A*, *STXBP1*, and *SYNGAP1.* To support exploration of their unique trajectories and outcomes, we generated annotated datasets encompassing 97,256 annotated clinical concepts representing 3,424 years of patient data distributed across 466 individuals.

### Longitudinal phenotypes derived from RWD replicate established disease trajectories

Given the importance of early recognition and work-up by clinicians in shortening the diagnostic odyssey, we employed enrichment analysis of annotated clinical concepts to differentiate between the six DEEs in infancy and childhood. A distinct phenotypic gestalt was observed for FOXG1 syndrome, emerging within the first year of life. Individuals showed enrichment for core features of FOXG1 syndrome, including microcephaly, failure to thrive, and strabismus. By six years of age, clinical concepts representing the previously described hyperkinetic-dyskinetic movement disorder were enriched for, including dystonia and chorea.^30^ In individuals with variants in *KCNQ2*, *SCN8A*, and *STXBP1*, an early-onset epilepsy phenotype is apparent. This finding is consistent with what has been described previously,^27, 38, 39^ and is further supported through our evaluation of epilepsy burden using a novel composite severity metric.

More recently, the epilepsy phenotype for STXBP1-related disorder has been well characterized, with authors highlighting variability in seizure onset and semiology, as well as in observed electroencephalogram abnormalities.^17, 39^ Our findings modeling the seizure trajectory in a larger cohort of 85 individuals replicate those reported by Xian *et al.* (n=62), where seizure burden is highest in the first year of life with marked improvement observed in early childhood.^17^ Further, no single seizure type was enriched for in this cohort; occurrence of both focal and generalized seizure types were documented, providing additional support for phenotypic heterogeneity. We also identified enrichment for tremor and dysmetria in early life; coordination problems have been previously described in *STXBP1*-related disorder.^17, 39^ By replicating previously described phenotypes across the six DEEs evaluated here, we provide support for the validity of our approach in generating insights from RWD for rare disorders.

### Semantic similarity analysis highlights phenotypic homogeneity or heterogeneity

In this study, we relied on annotation of clinical concepts using a controlled ontology under the guidance of a consistent data modeling framework. In doing so, we enable computational analyses to manipulate large datasets in order to identify patterns within and between cohorts. For clinical trial designs that rely on use of retrospective natural history data for an external control, predictability in disease presentation and progression has been highlighted as a prerequisite.^40^ To investigate consistency within cohorts, we employed semantic similarity analysis to generate scores proportional to phenotypic similarity. The greatest scores amongst individuals with the same genetic etiology were observed for *FOXG1* and *KCNQ2*, whereas the lowest scores were observed for *STXBP1*. In support, Lewis-Smith *et al.* highlighted *KCNQ2* (n=8) as a gene with significant phenotypic homogeneity,^12^ which is in contrast to earlier work where *STXBP1* (n=14) was noted to have significant phenotypic similarity, and *KCNQ2* (n=9) was associated with relatively limited phenotypic similarity.^16^ These discrepancies may reflect improved ability to characterize cohorts as a result of increased sample size and consistent data capture. Our findings suggest that FOXG1 syndrome and KCNQ2-related disorder may be well-suited for externally-controlled clinical trials, given the importance of limiting cohort variability.^40^

### Technical and operational protocols can address RWD quality concerns

The RWD platform described here was developed to accelerate collection of longitudinal phenotypic data to inform and optimize clinical trial design in rare disorders. Patient-mediated platforms can facilitate comprehensive data generation through longitudinal collection of medical records across all sites of care. In our approach, we further prepare clinical data for research through systematic annotation performed by a centralized team. Annotated data are fit to a data model which was designed to support endpoint identification and characterization. The scope of data capture spans 18 clinical data entities, including physical exam findings, hospitalizations, seizure frequency, developmental outcomes, and medications, and represents an expansion beyond prior efforts.^12, 13, 16^ This allows for increased granularity and richness of the resulting dataset. In addition, by establishing a remote design that is agnostic to clinical sites, we increased patient access to research and reduced likelihood of attrition by minimizing burden to participants. In aggregate, we demonstrate the platform’s ability to generate rich longitudinal datasets for representative cohorts to improve understanding of the phenotypic spectrum in rare disorders. As further validation, a subset of the data presented here for *SCN2A* was used to support an Investigational New Drug application for an antisense oligonucleotide targeting gain-of-function variants.^41^

The use of RWD to describe natural history, particularly that derived from EHRs, represents a secondary application of medical information that is not systematically generated for research purposes. These concerns are further compounded by variability in data sources and a lack of standardization for the documentation or evaluation of data quality. In response to these concerns, the Data Quality Harmonization Framework (DQHF) was developed to define axes by which the quality of EHR-based datasets can be measured.^42^ This framework has been successfully employed by the ‘All of Us’ Research Program Clinical Data Research Network, where data from multiple clinical sites are centralized.^43^ The axes defined by DQHF include conformance, completeness, and plausibility. We have implemented technical and operational processes to maximize quality across each of the three axes. Conformance measures a dataset’s compliance with internal and/or external formatting requirements; data generated via the Ciitizen platform is constrained by a data model and by the use of standard ontologies. Where appropriate, validation checkpoints have been implemented to further restrict generation of non-conforming data. To address data completeness, we use a combined approach: (i) collected medical records are systematically reviewed for completeness against a standard definition, (ii) the data model supports redundant annotation to highlight and supplement possible missingness, and (iii) generated data undergoes review by advanced practice providers with relevant training and clinical experience. To achieve plausibility, a measure of data accuracy or fidelity, extraction from high quality sources is prioritized and where indicated, multiple pieces of evidence are used to support annotated data elements. In support, the data presented here reflect established disease trajectories where available, and age-dependent phenotypes emerge at appropriate times, which represents a previously proposed quality control measure.^13^

### Application of RWD to clinical trial development and design for DEEs

The detailed phenotyping performed here across large populations supports identification and characterization of primary and secondary endpoints. For example, by combining enrichment analysis with relative frequency of propagated clinical concepts, we highlight candidate phenotypes by which to measure treatment efficacy in a trial setting. For individuals with a variant in *SYNGAP1*, both communication disorder and speech delay are enriched for and are frequently diagnosed. Therefore, use of a validated measure of communication, like the Observer-Reported Communication Ability (ORCA) measure, may represent an appropriate candidate endpoint. Similarly, enrichment was observed for behavioral challenges and neuropsychological diagnoses in the setting of universal developmental differences. Standardized assessments or historical data describing neurodevelopmental outcomes may be successful measures of efficacy. Finally, seizures were frequently reported in the cohort (83.1%) with specific enrichment for generalized seizure types, like absence and atonic seizures. By overlaying findings from the novel seizure severity score presented here, use of generalized seizures as an endpoint would be most successful in participants aged 2-6 years where epilepsy disease burden is highest.

With established frameworks by which to evaluate RWD quality, and demonstrated efficacy in describing longitudinal phenotypes, opportunities arise to expand the application of RWD beyond preclinical studies. Although prospective natural history studies remain a gold standard in rare disorders, the FDA and other regulatory agencies acknowledge that these studies are not always feasible or ethical; this is particularly true for rare disorders associated with significant morbidity or mortality,^44^ and is exacerbated in small, geographically distributed populations. The FDA has accepted a variety of data sources as supporting evidence or external controls in clinical trials for rare disorders, including data sourced from patient medical records.^40^ In the appropriate settings, RWD can improve efficiency and cost-effectiveness in developing and implementing clinical trials.^45^ The platform described here highlights a scalable approach to maximize quality and versatility of RWD in the characterization of rare disorders.

Ongoing expansion and harmonization of RWD sources can further enable drug development for rare disorders. For example, pairing retrospective EHR data with patient-generated health data prospectively collected via surveys or validated measures can strengthen understanding of the lived experience for patients and caregivers. Collection of electrophysiology and imaging studies allows for systematic characterization by a central reviewer, as well as identification of putative biomarkers. Further, additional data sources can be used to validate events described in medical records. Tokenization, the process of substituting sensitive descriptors with non-sensitive surrogates, can link participants enrolled in Ciitizen with claims databases to verify use of medications, or evaluate the potential for missing data. With the appropriate consents and/or protocols in place, tokenization also enables sharing across platforms, addressing the siloing of data that can impede research progress for rare disorders. Most notably, the generation and application of RWD for rare disorders research is growing. As an example, since Ciitizen launched support for rare disorders in late 2020, we have connected with >3000 institutions and providers in the United States, annotated >400,000 clinical concepts, and generated data representing >10,000 patient years.

### Limitations

The primary limitation of our study lies in the use of EHR data which is generated for the purposes of documenting clinical care. Although we employed several measures to address data quality concerns, this represents an opportunistic data source that may include imperfect and inconsistent documentation. For example, although documentation of physical exam findings are often structured and systematic, clinical phenotypes and diagnoses are less so. In this work, absence of documentation of a clinical phenotype was interpreted to mean the phenotype was not present in the participant. Although comprehensiveness and redundancy of data capture are intended to reduce its likelihood, it is possible that a phenotype may exist for a participant who was not screened or described accordingly.

## Conclusion

As the promise of precision medicine becomes reality for a subset of rare disorders, scalable approaches to phenotypic characterization are critical in supporting clinical trial design and implementation. In this work, we describe the capabilities of a patient-mediated RWD platform in identifying candidate endpoints for use in clinical trials for six DEEs. By employing computational approaches that leverage the data’s structure and quality, we enable detailed characterization of phenotypes throughout development, highlighting the ability to differentiate between patients and cohorts. With appropriate study design and protocols to maximize data quality, RWD represents an opportunity to increase efficiency and cost-effectiveness of natural history data collection, particularly in rare disorder populations with high unmet needs.

## Data Availability

The raw data that support the analyses presented here contain sensitive and protected health information for participants and is therefore not openly available. Requests for data access can be made to, where we can confirm the proposed research scope is consistent with platform consent language and is reviewed and/or approved by an institutional review board.

## Acknowledgements

We extend our gratitude to the participants and their families, as well as to our partnered advocacy groups for recruitment support: FOXG1 Research Foundation, KCNQ2 Cure Alliance, FamilieSCN2A, Cute Syndrome Foundation, International SCN8A Alliance, STXBP1 Foundation, Shay Emma Hammer Research Foundation, and SynGAP Research Fund. We also highlight Data Curation performed by the Ciitizen Operations team.

## Author Contributions

EB: Conceptualization, Formal Analysis, Methodology, Supervision, Visualization, Writing - Original Draft, Writing - Review & Editing; JK and RLM: Conceptualization, Formal Analysis, Methodology, Software, Visualization, Writing - Review & Editing; DM: Conceptualization, Writing - Review & Editing; AMBL: Conceptualization, Formal Analysis, Methodology, Software, Supervision, Visualization, Writing - Original Draft, Writing - Review & Editing.

## Declaration of Interests

All authors are current or former employees and shareholders of Invitae, which owns the medical record data extraction platform, Ciitizen.

## Notes

### Competing Interest Statement

All authors are current or former employees and shareholders of Invitae, Corp, which owns the medical record data extraction platform, Ciitizen.

### Funding Statement

This study was funded by Invitae, Corp.

### Author Declarations

Pearl IRB, an independent IRB, gave ethical approval for this work.

### Summary of Updates

Updated spacing between figure 2 legend and main text.

## References

1. Scheffer, I.E., Berkovic, S., Capovilla, G., Connolly, M.B., French, J., Guilhoto, L., Hirsch, E., Jain, S., Mathern, G.W., Moshé, S.L., et al. (2017). ILAE classification of the epilepsies: Position paper of the ILAE Commission for Classification and Terminology. Epilepsia. 58, 512–521. 10.1111/epi.13709.

2. Han, Z., Chen, C., Christiansen, A., Ji, S., Lin, Q., Anumonwo, C., Liu, C., Leiser, S.C., Meena, Aznarez, I., et al. (2020). Antisense oligonucleotides increase Scn1a expression and reduce seizures and SUDEP incidence in a mouse model of Dravet syndrome. Sci. Transl. Med. 12, eaaz6100. 10.1126/scitranslmed.aaz6100.

3. Li, M., Jancovski, N., Jafar-Nejad, P., Burbano, L.E., Rollo, B., Richards, K., Drew, L., Sedo, A., Heighway, J., Pachernegg, S., et al. (2021). Antisense oligonucleotide therapy reduces seizures and extends life span in an SCN2A gain-of-function epilepsy model. J. Clin. Invest. 131, e152079. 10.1172/JCI152079.

4. Milazzo, C., Mientjes, E.J., Wallaard, I., Rasmussen, S.V., Erichsen, K.D., Kakunuri, T., van der Sman, A.S.E., Kremer, T., Miller, M.T., Hoener, M.C., et al. (2021). Antisense oligonucleotide treatment rescues UBE3A expression and multiple phenotypes of an Angelman syndrome mouse model. JCI Insight 6, e145991. 10.1172/jci.insight.145991.

5. Helbig, I., and Tayoun, A.N.A. (2016). Understanding Genotypes and Phenotypes in Epileptic Encephalopathies. Mol. Syndromol. 7, 172–181. 10.1159/000448530.

6. US Food and Drug Administration (2018). Framework for FDA’s Real-World Evidence Program.

7. Purpura, C.A., Garry, E.M., Honig, N., Case, A., and Rassen, J.A. (2022). The Role of Real-World Evidence in FDA-Approved New Drug and Biologics License Applications. Clin. Pharmacol. Ther. 111, 135–144. 10.1002/cpt.2474.

8. Naumann-Winter, F., Wolter, F., Hermes, U., Malikova, E., Lilienthal, N., Meier, T., Kalland, M.E., and Magrelli, A. (2022). Licensing of Orphan Medicinal Products-Use of Real-World Data and Other External Data on Efficacy Aspects in Marketing Authorization Applications Concluded at the European Medicines Agency Between 2019 and 2021. Front. Pharmacol. 13, 920336. 10.3389/fphar.2022.920336.

9. Tisdale, A., Cutillo, C.M., Nathan, R., Russo, P., Laraway, B., Haendel, M., Nowak, D., Hasche, C., Chan, C.-H., Griese, E., et al. (2021). The IDeaS initiative: pilot study to assess the impact of rare diseases on patients and healthcare systems. Orphanet J. Rare Dis. 16, 429. 10.1186/s13023-021-02061-3.

10. Banda, J.M., Seneviratne, M., Hernandez-Boussard, T., and Shah, N.H. (2018). Advances in Electronic Phenotyping: From Rule-Based Definitions to Machine Learning Models. Annu. Rev. Biomed. Data Sci. 1, 53–68. 10.1146/annurev-biodatasci-080917-013315.

11. Köhler, S., Øien, N.C., Buske, O.J., Groza, T., Jacobsen, J.O.B., McNamara, C., Vasilevsky, N., Carmody, L.C., Gourdine, J.P., Gargano, M., et al. (2019). Encoding Clinical Data with the Human Phenotype Ontology for Computational Differential Diagnostics. Curr. Protoc. Hum. Genet. 103, e92. 10.1002/cphg.92.

12. Lewis-Smith, D., Ganesan, S., Galer, P.D., Helbig, K.L., McKeown, S.E., O’Brien, M., Khankhanian, P., Kaufman, M.C., Gonzalez, A.K., Felmeister, A.S., et al. (2021). Phenotypic homogeneity in childhood epilepsies evolves in gene-specific patterns across 3251 patient-years of clinical data. Eur. J. Hum. Genet. 29, 1690–1700. 10.1038/s41431-021-00908-8.

13. Ganesan, S., Galer, P.D., Helbig, K.L., McKeown, S.E., O’Brien, M., Gonzalez, A.K., Felmeister, A.S., Khankhanian, P., Ellis, C.A., and Helbig, I. (2020). A longitudinal footprint of genetic epilepsies using automated electronic medical record interpretation. Genet. Med. 22, 2060–2070. 10.1038/s41436-020-0923-1.

14. Helbig, I., Lopez-Hernandez, T., Shor, O., Galer, P., Ganesan, S., Pendziwiat, M., Rademacher, A., Ellis, C.A., Hümpfer, N., Schwarz, N., et al. (2019). A Recurrent Missense Variant in AP2M1 Impairs Clathrin-Mediated Endocytosis and Causes Developmental and Epileptic Encephalopathy. Am. J. Hum. Genet. 104, 1060–1072. 10.1016/j.ajhg.2019.04.001.

15. Crawford, K., Xian, J., Helbig, K.L., Galer, P.D., Parthasarathy, S., Lewis-Smith, D., Kaufman, M.C., Fitch, E., Ganesan, S., O’Brien, M., et al. (2021). Computational analysis of 10,860 phenotypic annotations in individuals with SCN2A-related disorders. Genet. Med. 23, 1263–1272. 10.1038/s41436-021-01120-1.

16. Galer, P.D., Ganesan, S., Lewis-Smith, D., McKeown, S.E., Pendziwiat, M., Helbig, K.L., Ellis, C.A., Rademacher, A., Smith, L., Poduri, A., et al. (2020). Semantic Similarity Analysis Reveals Robust Gene-Disease Relationships in Developmental and Epileptic Encephalopathies. Am. J. Hum. Genet. 107, 683–697. 10.1016/j.ajhg.2020.08.003.

17. Xian, J., Parthasarathy, S., Ruggiero, S.M., Balagura, G., Fitch, E., Helbig, K., Gan, J., Ganesan, S., Kaufman, M.C., Ellis, C.A., et al. (2021). Assessing the landscape of STXBP1-related disorders in 534 individuals. Brain. 145, 1668–1683. 10.1093/brain/awab327.

18. Son, J.H., Xie, G., Yuan, C., Ena, L., Li, Z., Goldstein, A., Huang, L., Wang, L., Shen, F., Liu, H., et al. (2018). Deep Phenotyping on Electronic Health Records Facilitates Genetic Diagnosis by Clinical Exomes. Am. J. Hum. Genet. 103, 58–73. 10.1016/j.ajhg.2018.05.010.

19. Richards, S., Aziz, N., Bale, S., Bick, D., Das, S., Gastier-Foster, J., Grody, W.W., Hegde, M., Lyon, E., Spector, E., et al. (2015). Standards and guidelines for the interpretation of sequence variants: a joint consensus recommendation of the American College of Medical Genetics and Genomics and the Association for Molecular Pathology. Genet. Med. 17, 405–424. 10.1038/gim.2015.30.

20. Nottke, A., Alan, S., Brimble, E., Cardillo, A.B., Henderson, L., Littleford, H.E., Rojahn, S., Sage, H., Taylor, J., West-Odell, L.E., et al. (2023). Validation and clinical discovery demonstration of a real-world data extraction platform. medRxiv. 2023.02.21.23286092. 10.1101/2023.02.21.23286092.

21. Terminologies-systems - FHIR v5.0.0-cibuild https://build.fhir.org/terminologies-systems.html.

22. SNOMED. https://www.snomed.org/.

23. Nelson, S.J., Zeng, K., Kilbourne, J., Powell, T., and Moore, R. (2011). Normalized names for clinical drugs: RxNorm at 6 years. J. Am. Med. Inform. Assoc. 18, 441–448. 10.1136/amiajnl-2011-000116.

24. Resnik, P. (1995). Using Information Content to Evaluate Semantic Similarity in a Taxonomy. 10.48550/arXiv.cmp-lg/9511007.

25. Köhler, S., Schulz, M.H., Krawitz, P., Bauer, S., Dölken, S., Ott, C.E., Mundlos, C., Horn, D., Mundlos, S., and Robinson, P.N. (2009). Clinical Diagnostics in Human Genetics with Semantic Similarity Searches in Ontologies. Am. J. Hum. Genet. 85, 457–464. 10.1016/j.ajhg.2009.09.003.

26. Fitzgerald, M.P., Kaufman, M.C., Massey, S.L., Fridinger, S., Prelack, M., Ellis, C., Ortiz-Gonzalez, X., Fried, L.E., DiGiovine, M.P., Collaborative, C.P.E.P., et al. (2021). Assessing seizure burden in pediatric epilepsy using an electronic medical record–based tool through a common data element approach. Epilepsia. 62, 1617–1628. 10.1111/epi.16934.

27. Malerba, F., Alberini, G., Balagura, G., Marchese, F., Amadori, E., Riva, A., Vari, M.S., Gennaro, E., Madia, F., Salpietro, V., et al. (2020). Genotype-phenotype correlations in patients with de novo KCNQ2 pathogenic variants. Neurol. Genet. 6, e528. 10.1212/NXG.0000000000000528.

28. Knowles, J.K., Helbig, I., Metcalf, C.S., Lubbers, L.S., Isom, L.L., Demarest, S., Goldberg, E.M., George, A.L., Lerche, H., Weckhuysen, S., et al. (2022). Precision medicine for genetic epilepsy on the horizon: Recent advances, present challenges, and suggestions for continued progress. Epilepsia. 63, 2461–2475. 10.1111/epi.17332.

29. Cellini, E., Vignoli, A., Pisano, T., Falchi, M., Molinaro, A., Accorsi, P., Bontacchio, A., Pinelli, L., Giordano, L., Guerrini, R., et al. (2016). The hyperkinetic movement disorder of FOXG1-related epileptic-dyskinetic encephalopathy. Dev. Med. Child Neurol. 58, 93–97. 10.1111/dmcn.12894.

30. Papandreou, A., Schneider, R.B., Augustine, E.F., Ng, J., Mankad, K., Meyer, E., McTague, A., Ngoh, A., Hemingway, C., Robinson, R., et al. (2016). Delineation of the movement disorders associated with FOXG1 mutations. Neurology. 86, 1794–1800. 10.1212/WNL.0000000000002585.

31. Vegas, N., Cavallin, M., Maillard, C., Boddaert, N., Toulouse, J., Schaefer, E., Lerman-Sagie, T., Lev, D., Magalie, B., Moutton, S., et al. (2018). Delineating FOXG1 syndrome: From congenital microcephaly to hyperkinetic encephalopathy. Neurol. Genet. 4, e281. 10.1212/NXG.0000000000000281.

32. Reynolds, C., King, M.D., and Gorman, K.M. (2020). The phenotypic spectrum of SCN2A-related epilepsy. Eur. J. Paediatr. Neurol. EJPN Off. J. Eur. Paediatr. Neurol. Soc. 24, 117–122. 10.1016/j.ejpn.2019.12.016.

33. Shellhaas, R.A., Wusthoff, C.J., Tsuchida, T.N., Glass, H.C., Chu, C.J., Massey, S.L., Soul, J.S., Wiwattanadittakun, N., Abend, N.S., Cilio, M.R., et al. (2017). Profile of neonatal epilepsies: Characteristics of a prospective US cohort. Neurology. 89, 893–899. 10.1212/WNL.0000000000004284.

34. Berg, A.T., Coryell, J., Saneto, R.P., Grinspan, Z.M., Alexander, J.J., Kekis, M., Sullivan, J.E., Wirrell, E.C., Shellhaas, R.A., Mytinger, J.R., et al. (2017). Early-Life Epilepsies and the Emerging Role of Genetic Testing. JAMA Pediatr. 171, 863–871. 10.1001/jamapediatrics.2017.1743.

35. Shoichet, S.A., Kunde, S.-A., Viertel, P., Schell-Apacik, C., von Voss, H., Tommerup, N., Ropers, H.-H., and Kalscheuer, V.M. (2005). Haploinsufficiency of novel FOXG1B variants in a patient with severe mental retardation, brain malformations and microcephaly. Hum. Genet. 117, 536–544. 10.1007/s00439-005-1310-3.

36. Seltzer, L.E., Ma, M., Ahmed, S., Bertrand, M., Dobyns, W.B., Wheless, J., and Paciorkowski, A.R. (2014). Epilepsy and outcome in FOXG1-related disorders. Epilepsia. 55, 1292–1300. 10.1111/epi.12648.

37. Mitter, D., Pringsheim, M., Kaulisch, M., Plümacher, K.S., Schröder, S., Warthemann, R., Abou Jamra, R., Baethmann, M., Bast, T., Büttel, H.-M., et al. (2018). FOXG1 syndrome: genotype-phenotype association in 83 patients with FOXG1 variants. Genet. Med. 20, 98–108. 10.1038/gim.2017.75.

38. Johannesen, K.M., Liu, Y., Koko, M., Gjerulfsen, C.E., Sonnenberg, L., Schubert, J., Fenger, C.D., Eltokhi, A., Rannap, M., Koch, N.A., et al. (2022). Genotype-phenotype correlations in SCN8A-related disorders reveal prognostic and therapeutic implications. Brain. 145, 2991–3009. 10.1093/brain/awab321.

39. Balagura, G., Xian, J., Riva, A., Marchese, F., Ben Zeev, B., Rios, L., Sirsi, D., Accorsi, P., Amadori, E., Astrea, G., et al. (2022). Epilepsy Course and Developmental Trajectories in STXBP1-DEE. Neurol. Genet. 8, e676. 10.1212/NXG.0000000000000676.

40. Jahanshahi, M., Gregg, K., Davis, G., Ndu, A., Miller, V., Vockley, J., Ollivier, C., Franolic, T., and Sakai, S. (2021). The Use of External Controls in FDA Regulatory Decision Making. Ther. Innov. Regul. Sci. 55, 1019–1035. 10.1007/s43441-021-00302-y.

41. Invitae’s Real-World Ciitizen Data Utilized in Praxis Precision Medicines’ PRAX-222 IND Filing https://ir.invitae.com/news-and-events/press-releases/press-release-details/2022/Invitaes-Real-World-Ciitizen-Data-Utilized-in-Praxis-Precision-Medicines-PRAX-222-IND-Filing/default.aspx.

42. Kahn, M.G., Callahan, T.J., Barnard, J., Bauck, A.E., Brown, J., Davidson, B.N., Estiri, H., Goerg, C., Holve, E., Johnson, S.G., et al. (2016). A Harmonized Data Quality Assessment Terminology and Framework for the Secondary Use of Electronic Health Record Data. EGEMs. 4, 18. 10.13063/2327-9214.1244.

43. Engel, N., Wang, H., Jiang, X., Lau, C.Y., Patterson, J., Acharya, N., Beaton, M., Sulieman, L., Pavinkurve, N., and Natarajan, K. (2022). EHR Data Quality Assessment Tools and Issue Reporting Workflows for the “All of Us” Research Program Clinical Data Research Network. AMIA. Annu. Symp. Proc. 2022, 186–195.

44. U.S. Department of Health and Human Services, Food and Drug Administration, Center for Drug Evaluation and Research (CDER), and Center for Biologics Evaluation and Research (CBER) (2019). Rare Diseases: Common Issues in Drug Development: Guidance for Industry. Rare Dis.

45. Liu, J., Barrett, J.S., Leonardi, E.T., Lee, L., Roychoudhury, S., Chen, Y., and Trifillis, P. (2022). Natural History and Real-World Data in Rare Diseases: Applications, Limitations, and Future Perspectives. J. Clin. Pharmacol. 62, S38–S55. 10.1002/jcph.2134.

