## Supplemental Tables and Figures for "Computation of longitudinal phenotypes in 466 individuals with a developmental and epileptic encephalopathy enables clinical trial readiness"

### Supplementary Figures and Tables

#### Supplementary Figures

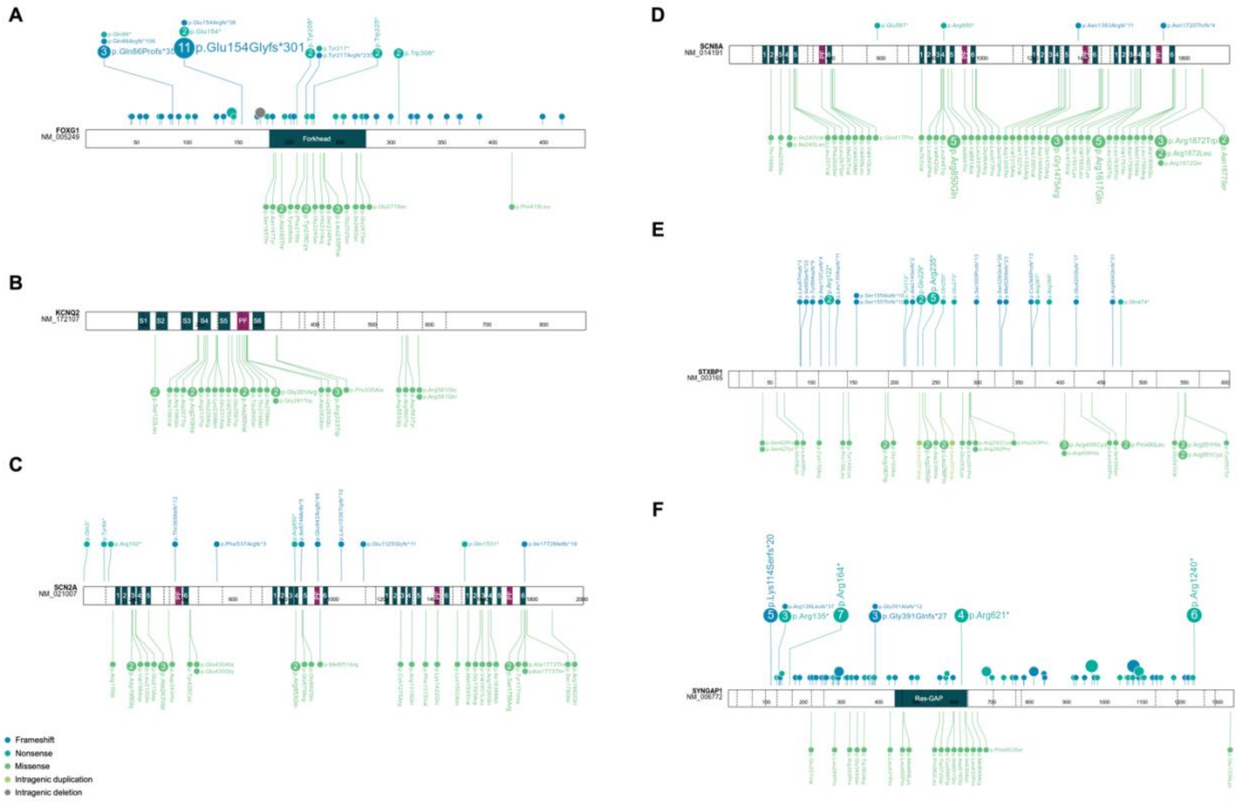


**Supplementary Figure 1. Distribution of sequence variants across six DEE cohorts**

The distribution of reported sequence variants is shown across the protein and relevant domains for FOXG1 **(A)**, KCNQ2 **(B)**, SCN2A **(C)**, SCN8A **(D)**, STXBP1 **(E)**, SYNGAP1 **(F)**. Recurrent variants are indicated by numbered circles. Forkhead = Forkhead DNA binding domain; S1-S6 = transmembrane segments; PF = pore-forming domain. Exons are separated by dotted vertical lines.

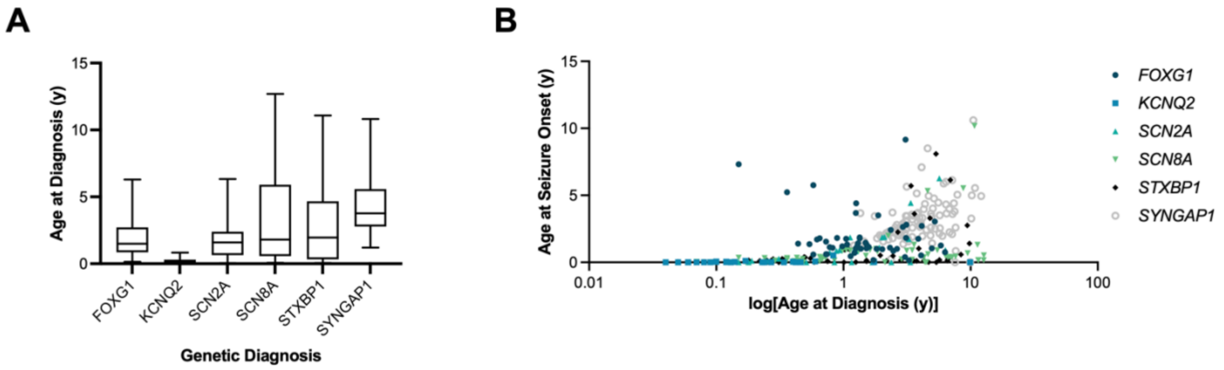


**Supplementary Figure 2. Age at genetic diagnosis and correlation with age at seizure onset**

The distribution of age at diagnosis is shown for each DEE cohort with outliers excluded **(A)**, in addition to the correlation between age at diagnosis and age at seizure onset in years **(B)**. The $R2^2$ values for each cohort are as follows: *FOXG1* = 0.0064, *KCNQ2* = 0.0000, *SCN2A* = 0.4142, *SCN8A* = 0.1337, *STXBP1* = 0.1995, *SYNGAP1* = 0.1661.

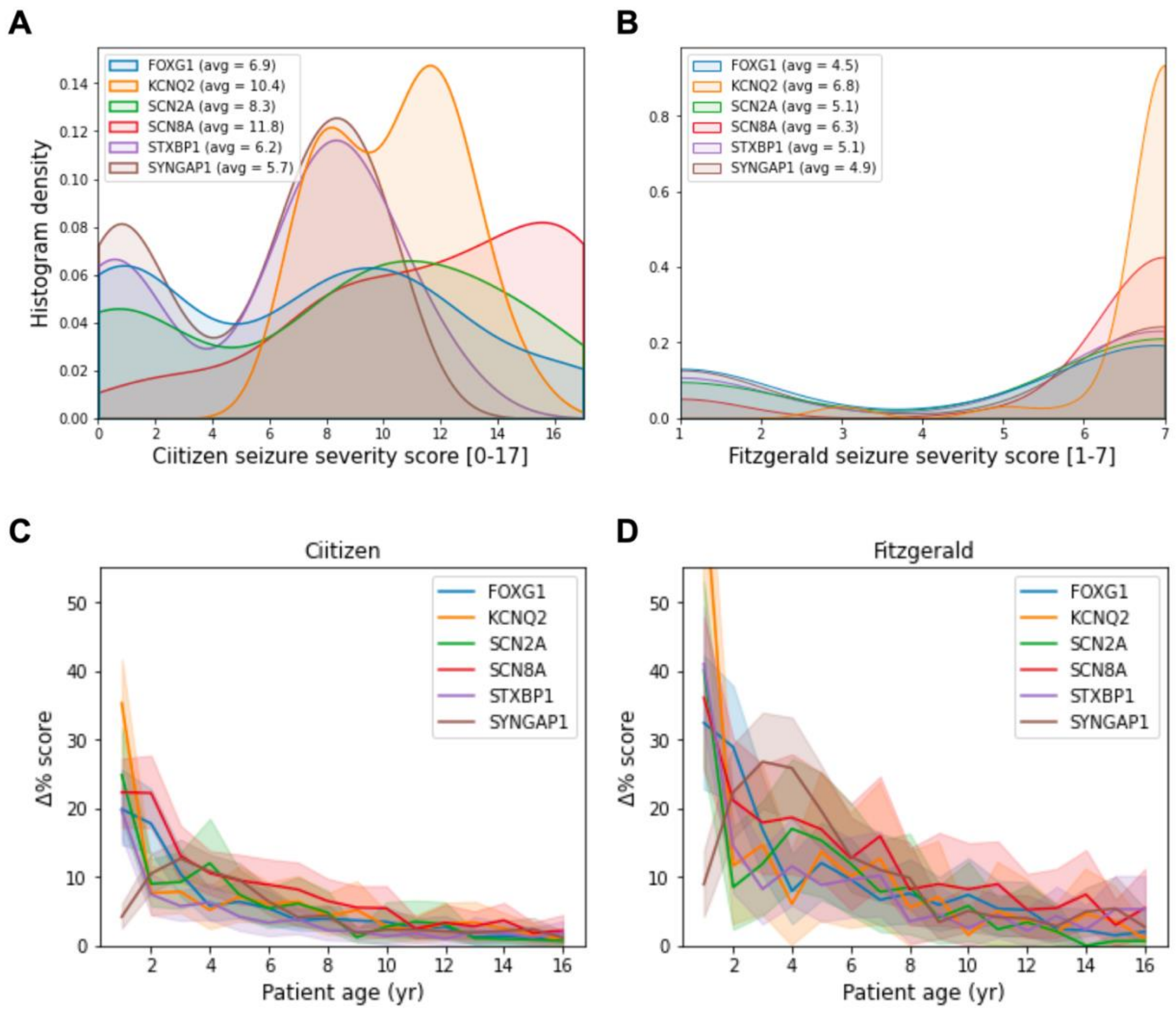


**Supplementary Figure 3. Evaluating a novel composite seizure severity metric**

Relative frequency of maximum seizure severity scores are shown for the six DEE cohorts, with higher severity scores indicating increased burden of epilepsy **(A)**. The performance of the composite severity metric described here was compared to a previously published score[26] **(B)**. To evaluate robustness of the composite metric, we plotted natural variation in scoring (as a result of age, change in management, etc.) **(C, D)**. Average change in score by year of age is shown with 95% confidence intervals.

#### Supplementary Tables

##### Supplementary Table 1. Scope and structure of data capture

| Clinical Data Entity | Field | Example | Example Code | Date(s) Included | Terminology |
|---|---|---|---|---|---|
| adverse effects | adverse effect | Fatigue | 84229001 | Y | SNOMED CT |
| adverse effects | associated medication or procedure | Clobazam | 21241 | | RXNORM SNOMED CT |
| clinical diagnosis | clinical diagnosis | Microcephaly | 1148757008 | Y | SNOMED CT |
| demographics | age at diagnosis | 1 | | | |
| demographics | year of birth | 2010 | | Y | |
| demographics | sex | Female | | | |
| development | domain | Gross Motor Development | | | |
| development | milestone | Ability to sit unsupported | 302039004 | | SNOMED CT |
| development | status | Unable | 371151008 | Y | SNOMED CT |
| devices | domain | Language Development | | | |
| devices | device | Eye gaze device | 15161000224107 | Y | SNOMED CT |
| diagnosis feature | diagnosis feature | Decreased | 1250004 | Y | SNOMED CT |
| diagnosis feature | associated diagnosis | Sleep disorder | 39898005 | | SNOMED CT |
| diagnostic procedures | procedure | Video EEG | 252738008 | Y | SNOMED CT |
| diagnostic procedures | procedure findings | Myoclonic seizure | 37356005 | | SNOMED CT |

| | | | | | |
|---|---|---|---|---|---|
| diagnostic procedures | procedure indication | Epilepsy | 84757009 | | SNOMED CT |
| exam findings | exam section | Coordination | | | |
| exam findings | exam finding | Incoordination | 281016006 | Y | SNOMED CT |
| genetics | test company | Baylor Genetics | | | |
| genetics | test name | Comprehensive Epilepsy Panel | | Y | |
| genetics | transcript ID | NM_005249 | | | |
| genetics | gene | FOXG1 | 3811 | | HGNC |
| genetics | variant DNA | c.701C>T | | | |
| genetics | variant protein | p.Ser234Phe | | | |
| genetics | inheritance | De novo (not inherited) | 13211000224104 | | SNOMED CT |
| genetics | interpretation | Pathogenic | 25141000224100 | | SNOMED CT |
| growth parameters | growth parameter | Body weight | 29463-7 | Y | LOINC |
| growth parameters | growth parameter value | 3.5 | | | |
| growth parameters | growth parameter unit | kg | | | UCUM |
| hospital admissions | admission diagnosis | Respiratory distress | 271825005 | Y | SNOMED CT |
| hospital admissions | significant event | Hospital acquired infection | 408678008 | | SNOMED CT |
| medications | medication | Valproate | 40254 | Y | RXNORM |
| medications | medication dose | 50 | | | |

| | | | | | |
|---|---|---|---|---|---|
| medications | medication unit | mg/8h | | | UCUM |
| medications | medication indication | Epilepsy | 84757009 | | SNOMED CT |
| menarche | age at menarche | 12 | | Y | SNOMED CT |
| primary diagnosis | primary diagnosis | FOXG1 Syndrome | 702450004 | Y | SNOMED CT |
| seizure history | seizure type | Febrile seizure | 432354000 | Y | SNOMED CT |
| seizure history | seizure value | 2 | | | |
| seizure history | seizure unit | Per week | | | |
| standardized assessment | standardized assessment | Capute scales | 12431000224103 | Y | SNOMED CT |
| standardized assessment | standardized assessment domain | CLAMS, Age Equivalent | | | |
| standardized assessment | standardized assessment value | 3.6 months | | | |
| therapeutic procedures | therapeutic procedure | Ketogenic diet | 765060000 | Y | SNOMED CT |
| laboratory studies | analyte | Valproate [Mass/volume] in Serum or Plasma | 4086-5 | Y | LOINC |
| laboratory studies | analyte value | 53 | | | |
| laboratory studies | analyte range from | 50 | | | |
| laboratory studies | analyte range to | 125 | | | |
| laboratory studies | analyte unit | ug/mL | | | UCUM |

**Supplementary Table 2. Domain structure of the composite seizure severity score**

| Domain | Scale | Scoring |
|---|---|---|
| Seizure frequency | 0-5 | 0 = seizure-free<br>1 = ≥1 seizure per year<br>2 = ≥1 seizure per month<br>3 = ≥1 seizure per week<br>4 = 1 seizure per day<br>5 = >1 seizure per day |
| Concomitant ASMs* | 0-5 | 0 = 0 ASMs used in the same time interval<br>1 = 1 ASM used in the same time interval<br>2 = 2 ASMs used in the same time interval<br>3 = 3 ASMs used in the same time interval<br>4 = 4 ASMs used in the same time interval<br>5 = 5+ ASMs used in the same time interval |
| Prolonged seizures (>5 minutes) | 0-2 | 0 = No prolonged seizures<br>1 = 1 prolonged seizure<br>2 = >1 prolonged seizure |
| Hospital admissions for prolonged seizure(s) and/or increased seizure frequency | 0-5 | 0 = No hospitalizations<br>1 = 1 hospitalization<br>2 = 2 hospitalizations<br>3 = 3 hospitalizations<br>4 = 4 hospitalizations<br>5 = 5+ hospitalizations |

Scoring rubric for the composite seizure severity score described in this study.

*Concomitant ASMs excludes prescription or use of rescue medications

**Supplementary Table 3. Data collection metrics for DEE cohorts**

| Cohort | mean # institutions (range) | mean # pages (range) | mean # patient data years (range) |
|---|---|---|---|
| *FOXG1* | 4.83 (1-21) | 1158.36 (66-8358) | 6.87 (0-27) |
| *KCNQ2* | 5.61 (1-17) | 2000.97 (282-14095) | 7.55 (1-23) |
| *SCN2A* | 7.37 (1-21) | 1417.47 (47-9178) | 6.80 (0-21) |
| *SCN8A* | 5.70 (1-20) | 2361.03 (117-16972) | 7.61 (0-23) |
| *STXBP1* | 6.11 (1-26) | 781.21 (63-7824) | 7.19 (1-24) |
| *SYNGAP1* | 5.93 (1-30) | 704.19 (33-5026) | 7.77 (1-48) |
| **All** | **5.84 (1-30)** | **1211.00 (33-16972)** | **7.35 (0-48)** |

Metrics describing the comprehensiveness of medical record collection by mean number of institutions, pages, and representative years of patient data.

**Supplementary Table 4. X-fold enrichment of clinical phenotypes in the first six years of life**

| diagnosis | Code | *FOXG1* | *KCNQ2* | *SCN2A* | *SCN8A* | *STXBP1* | *SYNGAP1* |
|---|---|---|---|---|---|---|---|
| microcephaly | 1148757008 | 11.093 | | | | | |
| dependency on feeding tube | 12991000224107 | | 2.734 | | | | |
| esotropia | 16596007 | 7.916 | | | | | |
| failure to thrive | 54840006 | 2.785 | | | | | |
| poor visual tracking | 14971000224102 | 2.558 | | | | | |
| dystonia | 15802004 | 2.742 | | | | | |
| strabismus | 22066006 | 4.011 | | | | | |
| appendicular hypertonia | 13291000224105 | | 3.007 | | | | |
| spasticity | 397790002 | 2.848 | | | | | |
| oropharyngeal dysphagia | 71457002 | 3.270 | | | | | |
| movement disorder | 60342002 | 3.899 | | | | | |
| respiratory distress | 271825005 | | 2.673 | | | | |
| chorea | 271700006 | 7.833 | | | | | |
| dependence on supplemental oxygen | 931000119107 | | 4.480 | | 2.897 | | |
| choreoathetosis | 43105007 | 8.356 | | | | | |
| abdominal pain | 21522001 | | | 3.161 | | | |
| amblyopia | 387742006 | 2.924 | | | | | |
| chronic cough | 68154008 | 3.481 | | | | | |
| seizure free | 370994008 | | 18.953 | | | | |
| altered mental status | 419284004 | | | | 3.248 | | |
| aggressive behavior | 61372001 | | | | | | 6.143 |
| apnea | 1023001 | | | | 7.146 | | |
| osteopenia | 312894000 | | | | 5.955 | | |
| tremor | 26079004 | | | | | 4.065 | |
| autism suspected | 401204006 | | | | | | 2.608 |
| sensory integration disorder | 425988004 | | | | | | 2.608 |
| dysmetria | 32566006 | | | | | 8.965 | |
| apraxia | 68345001 | | | | | | 4.563 |

| | | | | | | | |
|---|---|---|---|---|---|---|---|
| generalized epilepsy | 19598007 | | | | | | 5.476 |
| speech delay | 229721007 | | | | | | 2.825 |
| neonatal seizures | 87476004 | | | | | 5.603 | |

Clinical concepts shown here represent those enriched ≥2.5-fold in at least one of the six DEE cohorts, in the first six years of life.

**Supplementary Table 5. Percentage of individuals with altered seizure severity scores in response to data omissions**

| | **Fitzgerald *et al*. 2021[26]** | | **Composite Score** | |
|---|---|---|---|---|
| | **r = 10%** | **r = 20%** | **r = 10%** | **r = 20%** |
| *FOXG1* | 3.6% | 6.3% | 30% | 45% |
| *KCNQ2* | 2.5% | 4.5% | 33% | 56% |
| *SCN2A* | 3.7% | 5.0% | 33% | 46% |
| *SCN8A* | 3.1% | 2.8% | 34% | 47% |
| *STXBP1* | 3.2% | 4.5% | 31% | 44% |
| *SYNGAP1* | 6.8% | 7.5% | 26% | 37% |

The proportion of individuals whose scores are changed in response to 10% or 20% data omission.

**Supplementary Table 6. Magnitude of change in seizure severity scores for individuals with an altered score in response to data omissions**

| | **Fitzgerald *et al.* 2021[26]** | | **Composite Score** | |
|---|---|---|---|---|
| | **r = 10%** | **r = 20%** | **r = 10%** | **r = 20%** |
| *FOXG1* | 43% | 46% | 8% | 10% |
| *KCNQ2* | 43% | 54% | 7% | 9% |
| *SCN2A* | 98% | 98% | 9% | 11% |
| *SCN8A* | 44% | 39% | 8% | 9% |
| *STXBP1* | 68% | 75% | 8% | 8% |
| *SYNGAP1* | 87% | 83% | 11% | 10% |

The magnitude of change in seizure severity scores as a result of 10% or 20% data omissions.
